# Quality of life among adults with diabetes and hypertension in primary care outreach clinics in urban India: A cross-sectional study

**DOI:** 10.64898/2025.12.09.25341929

**Authors:** Swati Jha, Anoshmita Adhikary, Tejaswini Singh, Sonali Divate, Maninder Sethia

## Abstract

**Background:** Quality of life (QoL) is an important outcome for patients with diabetes and hypertension, influenced by disease control and sociodemographic factors. This study assessed QoL and examined its determinants among adults attending community-based primary care outreach clinics in urban slums of Mumbai, India.

**Methods:** A cross-sectional study involving 500 adults (≥18 years) categorized by disease status: diabetes mellitus (DM), hypertension (HT), both, or neither was conducted. QoL was assessed using the WHOQOL-BREF administered in local languages. Data on demographics, clinical variables, and QoL were collected electronically. Descriptive statistics, bivariate testing, and multivariable linear regression were performed.

**Results:** Mean age was 55.6 years (SD 13.5); 61% were female. Older age and female sex were significantly associated with lower QoL scores in physical, psychological, and environmental domains (p < 0.05). Poorly controlled DIABETES MELLITUS or HT correlated with significantly lower mental health–related QoL (p < 0.001). Multivariate analyses identified age, sex, income, smoking, disease status, and medication burden as independent QoL predictors.

**Conclusions:** QoL among adults with diabetes and hypertension in urban slums is negatively affected by older age, female gender, disease control, and socioeconomic factors. Integrated clinical and social interventions are vital to improve patient-centered outcomes.

## Introduction

Quality of life (QoL) captures individuals’ perceptions of their position in life relative to cultural and personal contexts and is a crucial outcome for managing chronic diseases such as diabetes and hypertension. Traditional health indicators measuring mortality and morbidity fail to capture the full impact of chronic conditions on daily functioning and well-being. Non-communicable diseases (NCDs) are now the leading causes of morbidity and mortality globally, posing major challenges to health systems and societies, particularly in low- and middle-income countries (LMICs).

Efficient primary care programs that promote prevention, early diagnosis, and chronic disease management are vital for addressing the growing burden of NCDs in urban poor communities. The Sir H. N. Reliance Foundation Hospital Health Outreach Program provides such care in Mumbai’s urban slums, focusing on diabetes and hypertension management.

While several studies have characterized QoL impairments associated with NCDs in India and elsewhere, there remains a knowledge gap regarding QoL among patients receiving treatment in community-based primary care outreach settings, especially among populations with multimorbidity. This study aims to fill that gap by assessing QoL and its determinants using the WHOQOL-BREF tool in this population.

Hypotheses tested include that age, sex, disease control, and socioeconomic factors significantly influence QoL across physical, psychological, social, and environmental domains among adults with diabetes and/or hypertension.

## Materials and Methods

### Study Design and Setting

This cross-sectional observational study was conducted between April 2024 and April 2025 in primary care outreach clinics of the Sir H. N. Reliance Foundation Hospital serving urban slum populations in Mumbai and Navi Mumbai.

### Participants

Adults aged 18 years and older registered with the outreach program and attending clinics during the study period were consecutively recruited. Participants were grouped by disease status: DIABETES MELLITUS only, HT only, both DIABETES MELLITUS and HT, or neither condition. Exclusion criteria included first-time visitors and those unable to provide informed consent due to cognitive impairment.

### Sample Size

Using prior QoL data, a calculated sample size of 423 was increased to 500 to ensure adequate subgroup power.

### Data Collection

The WHOQOL-BREF, a validated 26-item questionnaire assessing physical, psychological, social, and environmental QoL domains, was administered face-to-face in Hindi, Marathi, or Gujarati by trained social work staff using Google Forms. Additional data included demographics, income, education, lifestyle behaviours, disease status, duration, control, and medication use. Data collection was started on 11^th^ April 2025 and the recruitment ended on 5^th^ April 2025. Written informed consent was obtained by each participant in their mother tongue in front of a witness.

### Statistical Analysis

Data were analysed with STATA 17.0 and SPSS v22. Descriptive statistics summarized participant characteristics and QoL scores. Group comparisons used chi-square, t-tests, or non-parametric equivalents. Multivariable linear regression modelled associations between independent variables and domain-specific QoL scores adjusting for confounders. Statistical significance was set at p < 0.05.

### Ethics

Approval was obtained from the Institutional Ethics Committee of the Sir H. N. Reliance Foundation Hospital and Research Centre. Written informed consent was obtained from all participants.

## Results

### Participant Characteristics

Among 500 participants, the mean age was 55.6 (SD 13.5) years; 61% were female. Most (72.6%) had education ≤10th grade and 69.4% reported annual income ≤INR 85,000. Smoking was reported by 11.8%, and alcohol use was infrequent.

### Disease Profile

DIABETES MELLITUS prevalence was 41.2% with a median duration of 5 years. HYPERTENSION was present in 52.8% with median duration 3 years. Concomitant DIABETES MELLITUS and HYPERTENSION occurred in 31.2%. Approximately half reported controlled disease; the remainder reported uncontrolled or uncertain status. Median medication count was 3.

### QoL Scores and Bivariate Associations

Physical and psychological QoL scores declined with age (p < 0.001) and were lower among females (p < 0.01). Higher education and income correlated with better QoL scores across domains (p < 0.001). Participants with both DIABETES MELLITUS and HYPERTENSION had significantly lower QoL scores than those without these conditions (p < 0.001). Medication burden inversely correlated with QoL.

### Multivariable Analysis

Adjusted models confirmed older age and female sex as independent predictors of lower QoL across physical, psychological, and environmental domains. Higher income was positively associated with QoL scores. Smoking and multimorbidity predicted poorer psychological QoL, while disease control status affected mental health–related QoL substantially. Occasional alcohol consumption showed a paradoxical association with higher QoL scores.

**Table 1:** Baseline Characteristics of Study Participants (N=500)

| Parameter | N | Percentage (%) |
| --- | --- | --- |
| Age (years) |  |  |
| Mean (SD) | 55.6 (13.5) |  |
| 19-39 | 86 | 17.2 |
| 40-49 | 112 | 22.4 |
| 50-59 | 135 | 27.0 |
| 60-69 | 115 | 23.0 |
| $\geq 70$ | 52 | 10.4 |
| **Gender** |  |  |
| Male | 195 | 39.0 |
| Female | 305 | 61.0 |
| **Education** |  |  |
| Illiterate | 53 | 10.6 |
| Less than 10th | 181 | 36.2 |
| 10th | 129 | 25.8 |
| 12th | 72 | 14.4 |
| College | 44 | 8.8 |
| Professional | 21 | 4.2 |
| **Income (INR)** |  |  |
| $\leq 85,000$ | 243 | 69.4 |
| 85,000-160,000 | 76 | 21.7 |
| 160,000-270,000 | 12 | 3.4 |
| $\geq 270,000$ | 19 | 5.4 |
| **Smoking Status** |  |  |
| No | 426 | 85.4 |
| Yes | 59 | 11.8 |
| Occasionally | 14 | 2.8 |
| **Alcohol Consumption** |  |  |
| No | 470 | 94.0 |
| Yes | 8 | 1.6 |
| Occasionally | 22 | 4.4 |

**Table 2:** Disease Characteristics and Treatment Profile (N=500)

| Parameter | N | Percentage (%) |
| --- | --- | --- |
| **Diabetes Mellitus** |  |  |
| No | 294 | 58.8 |
| Yes | 206 | 41.2 |
| Duration of DM (years), Median (IQR) | 5 (2, 10) |  |
| **DM Control Status** (n=206) |  |  |
| No | 99 | 48.1 |
| Yes | 97 | 47.1 |
| Don't know | 10 | 4.9 |
| **Hypertension** |  |  |
| No | 236 | 47.2 |
| Yes | 264 | 52.8 |
| Duration of HT (years), Median (IQR) | 3 (1, 10) |  |
| **HT Control Status** (n=264) |  |  |
| No | 96 | 36.4 |
| Yes | 157 | 59.5 |
| Don't know | 11 | 4.2 |
| **Both DM and HT** |  |  |
| No | 307 | 68.8 |
| Yes | 139 | 31.2 |
| Duration of Both (years), Median (IQR) | 1 (3, 6) |  |
| **Both Conditions Control Status** (n=142) |  |  |
| No | 78 | 54.9 |
| Yes | 56 | 39.4 |
| Don't know | 8 | 5.6 |
| **Number of Medications** |  |  |
| Median (IQR) | 3 (1, 5) |  |

**Table 3:** Physical Domain Quality of Life Scores by Participant Characteristics.

| Variable | Mean | SD | p-value |
| --- | --- | --- | --- |
| **Overall** | 99.23 | 24.36 |  |
| **Age Groups** |  |  | <0.001 |
| 19-39 years | 116.47 | 22.38 |  |
| 40-49 years | 102.04 | 24.22 |  |
| 50-59 years | 99.70 | 20.71 |  |
| 60-69 years | 90.71 | 22.06 |  |
| ≥70 years | 82.31 | 22.48 |  |
| **Gender** |  |  | 0.0002 |
| Male | 104.27 | 23.30 |  |
| Female | 96.01 | 24.51 |  |
| **Education** |  |  | <0.001 |
| Illiterate | 84.38 | 20.52 |  |
| Less than 10th | 99.29 | 24.90 |  |
| 10th to 12th | 99.52 | 23.76 |  |
| College/Professional | 110.28 | 21.67 |  |
| **Marital Status** |  |  | 0.0001 |
| Never Married | 110.53 | 21.74 |  |
| Married | 99.83 | 23.61 |  |
| Separated/Widowed | 86.64 | 27.87 |  |
| **Income (INR)** |  |  | <0.001 |
| ≤85,000 | 94.60 | 21.07 |  |
| 85,000-160,000 | 109.89 | 21.32 |  |
| 160,000-270,000 | 114.67 | 22.19 |  |
| ≥270,000 | 104.63 | 19.88 |  |
| **Smoking** |  |  | 0.72 |
| No | 99.31 | 24.64 |  |
| Yes | 97.69 | 22.91 |  |
| Occasionally | 103.43 | 23.68 |  |
| **Alcohol** |  |  | 0.0001 |
| No | 98.30 | 24.27 |  |
| Yes | 96.00 | 14.81 |  |
| Occasionally | 120.36 | 19.55 |  |
| **Diabetes Mellitus** |  |  | <0.001 |
| No | 103.29 | 24.46 |  |
| Yes | 93.44 | 23.06 |  |
| **DM Duration (years)** |  |  | 0.22 |
| ≤2 | 97.18 | 22.00 |  |
| >2-5 | 93.00 | 24.52 |  |
| >5-10 | 93.12 | 19.91 |  |
| >10 | 87.70 | 25.56 |  |
| **DM Control** |  |  | 0.04 |
| No | 89.70 | 22.04 |  |
| Yes | 96.08 | 23.19 |  |
| Don't know | 104.80 | 26.72 |  |
| **Hypertension** |  |  | <0.001 |
| No | 105.71 | 24.19 |  |
| Yes | 93.44 | 23.05 |  |
| **HT Duration (years)** |  |  | 0.07 |
| ≤1 | 94.32 | 24.26 |  |
| >1-3 | 99.08 | 20.33 |  |
| >3-10 | 90.70 | 21.10 |  |
| >10 | 88.71 | 26.73 |  |
| **HT Control** |  |  | 0.29 |
| No | 91.50 | 25.96 |  |
| Yes | 93.99 | 21.39 |  |
| Don't know | 102.55 | 17.37 |  |
| **Both DM and HT** |  |  | <0.001 |
| No | 102.23 | 24.71 |  |
| Yes | 91.97 | 23.03 |  |
| **Both Duration (years)** |  |  | 0.07 |
| ≤1 | 94.32 | 24.26 |  |
| >1-3 | 99.08 | 20.33 |  |
| >3-6 | 91.14 | 20.63 |  |
| >6 | 89.40 | 24.49 |  |
| **Both Control** |  |  | 0.07 |
| No | 89.79 | 24.02 |  |
| Yes | 94.57 | 22.65 |  |
| Don't know | 109.00 | 16.25 |  |
| **Number of Medicines** |  |  | 0.005 |
| ≤1 | 99.11 | 25.64 |  |
| >1-3 | 97.77 | 21.81 |  |
| >3-5 | 91.88 | 21.80 |  |
| >5 | 88.21 | 22.04 |  |

**Table 4:** Psychological Domain Quality of Life Scores by Participant Characteristics.

| Variable | Mean | SD | p-value |
| --- | --- | --- | --- |
| **Overall** | 100.12 | 16.71 |  |
| **Age Groups** |  |  | <0.001 |
| 19-39 years | 107.63 | 13.96 |  |
| 40-49 years | 99.54 | 16.52 |  |
| 50-59 years | 100.59 | 15.35 |  |
| 60-69 years | 98.16 | 17.24 |  |
| ≥70 years | 92.08 | 18.92 |  |
| **Gender** |  |  | 0.004 |
| Male | 102.79 | 15.63 |  |
| Female | 98.41 | 17.17 |  |
| **Education** |  |  | <0.001 |
| Illiterate | 89.74 | 14.35 |  |
| Less than 10th | 100.18 | 14.66 |  |
| 10th to 12th | 101.03 | 18.19 |  |
| College/Professional | 105.60 | 15.76 |  |
| **Marital Status** |  |  | 0.004 |
| Never Married | 103.87 | 14.46 |  |
| Married | 100.67 | 16.12 |  |
| Separated/Widowed | 92.77 | 21.13 |  |
| **Income (INR)** |  |  | <0.001 |
| ≤85,000 | 96.86 | 13.90 |  |
| 85,000-160,000 | 106.21 | 15.18 |  |
| 160,000-270,000 | 110.00 | 8.44 |  |
| ≥270,000 | 104.84 | 13.10 |  |
| **Smoking** |  |  | 0.06 |
| No | 100.84 | 16.73 |  |
| Yes | 96.34 | 14.66 |  |
| Occasionally | 94.29 | 22.06 |  |
| **Alcohol** |  |  | 0.02 |
| No | 99.72 | 16.69 |  |
| Yes | 97.50 | 18.63 |  |
| Occasionally | 109.64 | 14.06 |  |
| **Diabetes Mellitus** |  |  | 0.005 |
| No | 101.86 | 16.46 |  |
| Yes | 97.63 | 16.79 |  |
| **DM Duration (years)** |  |  | 0.77 |
| ≤2 | 97.69 | 17.13 |  |
| >2-5 | 99.46 | 15.26 |  |
| >5-10 | 97.02 | 14.40 |  |
| >10 | 95.80 | 20.46 |  |
| **DM Control** |  |  | 0.05 |
| No | 95.39 | 17.37 |  |
| Yes | 98.89 | 16.33 |  |
| Don't know | 107.60 | 10.23 |  |
| **Hypertension** |  |  | 0.002 |
| No | 102.59 | 14.78 |  |
| Yes | 97.91 | 18.00 |  |
| **HT Duration (years)** |  |  | 0.30 |
| ≤1 | 99.95 | 17.39 |  |
| >1-3 | 99.88 | 16.84 |  |
| >3-10 | 95.95 | 17.80 |  |
| >10 | 95.20 | 20.67 |  |
| **HT Control** |  |  | 0.08 |
| No | 95.38 | 19.46 |  |
| Yes | 98.83 | 17.30 |  |
| Don't know | 106.91 | 9.65 |  |
| **Both DM and HT** |  |  | 0.005 |
| No | 101.73 | 16.29 |  |
| Yes | 96.83 | 17.98 |  |
| **Both Duration (years)** |  |  | 0.15 |
| ≤1 | 99.95 | 17.39 |  |
| >1-3 | 99.88 | 16.84 |  |
| >3-6 | 98.67 | 17.51 |  |
| >6 | 94.17 | 19.36 |  |
| **Both Control** |  |  | 0.06 |
| No | 95.18 | 18.10 |  |
| Yes | 98.07 | 18.13 |  |
| Don't know | 110.50 | 8.26 |  |
| **Number of Medicines** |  |  | 0.008 |
| ≤1 | 101.90 | 18.73 |  |
| >1-3 | 99.37 | 16.74 |  |
| >3-5 | 96.90 | 14.92 |  |
| >5 | 93.42 | 16.94 |  |

**Table 5:** Environmental Domain Quality of Life Scores by Participant Characteristics.

| Variable | Mean | SD | p-value |
| --- | --- | --- | --- |
| **Overall** | 126.22 | 20.98 |  |
| **Age Groups** |  |  | 0.0003 |
| 19-39 years | 134.70 | 19.66 |  |
| 40-49 years | 124.57 | 19.86 |  |
| 50-59 years | 126.93 | 19.07 |  |
| 60-69 years | 122.82 | 22.66 |  |
| ≥70 years | 121.46 | 22.89 |  |
| **Gender** |  |  | 0.30 |
| Male | 127.45 | 20.28 |  |
| Female | 125.44 | 21.41 |  |
| **Education** |  |  | <0.001 |
| Illiterate | 107.92 | 15.56 |  |
| Less than 10th | 125.19 | 19.65 |  |
| 10th to 12th | 129.59 | 20.91 |  |
| College/Professional | 133.60 | 20.30 |  |
| **Marital Status** |  |  | 0.73 |
| Never Married | 126.27 | 23.34 |  |
| Married | 125.97 | 20.56 |  |
| Separated/Widowed | 128.51 | 23.44 |  |
| **Income (INR)** |  |  | <0.001 |
| ≤85,000 | 115.52 | 17.70 |  |
| 85,000-160,000 | 136.32 | 17.46 |  |
| 160,000-270,000 | 144.67 | 11.67 |  |
| ≥270,000 | 133.47 | 16.40 |  |
| **Smoking** |  |  | 0.0002 |
| No | 127.59 | 20.79 |  |
| Yes | 115.80 | 19.66 |  |
| Occasionally | 129.71 | 21.55 |  |
| **Alcohol** |  |  | 0.29 |
| No | 126.13 | 20.90 |  |
| Yes | 118.00 | 19.36 |  |
| Occasionally | 131.27 | 22.98 |  |
| **Diabetes Mellitus** |  |  | 0.01 |
| No | 128.23 | 20.96 |  |
| Yes | 123.36 | 20.72 |  |
| **DM Duration (years)** |  |  | 0.79 |
| ≤2 | 122.54 | 20.43 |  |
| >2-5 | 125.38 | 21.86 |  |
| >5-10 | 121.40 | 19.79 |  |
| >10 | 124.30 | 21.19 |  |
| **DM Control** |  |  | 0.04 |
| No | 119.72 | 20.78 |  |
| Yes | 126.27 | 20.64 |  |
| Don't know | 131.20 | 15.18 |  |
| **Hypertension** |  |  | 0.04 |
| No | 128.24 | 20.06 |  |
| Yes | 124.42 | 21.65 |  |
| **HT Duration (years)** |  |  | 0.65 |
| ≤1 | 126.92 | 19.05 |  |
| >1-3 | 124.00 | 21.99 |  |
| >3-10 | 122.45 | 23.78 |  |
| >10 | 124.44 | 21.51 |  |
| **HT Control** |  |  | 0.049 |
| No | 120.67 | 23.15 |  |
| Yes | 126.04 | 20.84 |  |
| Don't know | 134.18 | 13.31 |  |
| **Both DM and HT** |  |  | 0.53 |
| No | 125.92 | 21.54 |  |
| Yes | 124.52 | 21.40 |  |
| **Both Duration (years)** |  |  | 0.66 |
| ≤1 | 126.92 | 19.05 |  |
| >1-3 | 124.00 | 21.99 |  |
| >3-6 | 124.38 | 24.48 |  |
| >6 | 122.55 | 22.21 |  |
| **Both Control** |  |  | 0.15 |
| No | 122.21 | 22.19 |  |
| Yes | 127.07 | 20.65 |  |
| Don't know | 135.50 | 12.73 |  |
| **Number of Medicines** |  |  | 0.0002 |
| ≤1 | 130.64 | 19.82 |  |
| >1-3 | 124.67 | 20.77 |  |
| >3-5 | 120.61 | 20.29 |  |
| >5 | 117.84 | 20.92 |  |

**Table 6.** Adjusted Multivariate Linear Regression Models for Quality of Life Domains Physical Domain Quality of Life.

| Variable | Coefficient | 95% CI | p-value | Interpretation |
| --- | --- | --- | --- | --- |
| **Disease Status** |  |  |  |  |
| Both DM & HT (vs. No) | -3.64 | (-7.65, 0.37) | 0.075 | Non-significant trend |
| **Age Group** |  |  |  |  |
| 19-39 years | Reference | - | - | - |
| 40-49 years | -13.22 | (-19.42, -7.02) | <0.001 | **Significant decrease** |
| 50-59 years | -14.13 | (-20.25, -8.01) | <0.001 | **Significant decrease** |
| 60-69 years | -22.36 | (-28.76, -15.96) | <0.001 | **Significant decrease** |
| ≥70 years | -30.35 | (-38.12, -22.59) | <0.001 | **Significant decrease** |
| **Gender** |  |  |  |  |
| Male | Reference | - | - | - |
| Female | -8.11 | (-12.42, -3.80) | <0.001 | **Significant decrease** |
| **Income (INR)** |  |  |  |  |
| <85,000 | Reference | - | - | - |
| 85,000-160,000 | 12.19 | (6.56, 17.82) | <0.001 | **Significant increase** |
| 160,000-270,000 | 11.29 | (-1.25, 23.83) | 0.078 | Non-significant trend |
| ≥270,000 | 7.68 | (-2.32, 17.68) | 0.132 | Non-significant |
| Missing | 6.88 | (2.27, 11.48) | 0.004 | **Significant increase** |
| **Marital Status** |  |  |  |  |
| Never Married | Reference | - | - | - |
| Married | -3.44 | (-11.78, 4.91) | 0.419 | Non-significant |
| Separated/Widowed | -9.26 | (-19.91, 1.39) | 0.088 | Non-significant trend |
| **Smoking** |  |  |  |  |
| No | Reference | - | - | - |
| Yes | -2.62 | (-9.07, 3.83) | 0.426 | Non-significant |
| Occasionally | -7.03 | (-18.90, 4.84) | 0.245 | Non-significant |
| **Alcohol** |  |  |  |  |
| No | Reference | - | - | - |
| Yes | -6.27 | (-21.96, 9.43) | 0.433 | Non-significant |
| Occasionally | 19.74 | (9.84, 29.64) | <0.001 | **Significant increase** |

**Table 7.** Psychological Domain Quality of Life.

| Variable | Coefficient | 95% CI | p-value | Interpretation |
| --- | --- | --- | --- | --- |
| **Disease Status** |  |  |  |  |
| Both DM & HT (vs. No) | -2.97 | (-5.92, -0.03) | 0.048 | **Significant decrease** |
| **Age Group** |  |  |  |  |
| 19-39 years | Reference | - | - | - |
| 40-49 years | -7.16 | (-11.72, -2.61) | 0.002 | **Significant decrease** |
| 50-59 years | -5.05 | (-9.54, -0.55) | 0.028 | **Significant decrease** |
| 60-69 years | -6.65 | (-11.35, -1.95) | 0.006 | **Significant decrease** |
| ≥70 years | -12.55 | (-18.26, -6.85) | <0.001 | **Significant decrease** |
| **Gender** |  |  |  |  |
| Male | Reference | - | - | - |
| Female | -5.15 | (-8.31, -1.98) | 0.001 | **Significant decrease** |
| **Income (INR)** |  |  |  |  |
| <85,000 | Reference | - | - | - |
| 85,000-160,000 | 8.48 | (4.34, 12.61) | <0.001 | **Significant increase** |
| 160,000-270,000 | 9.14 | (-0.07, 18.35) | 0.052 | Non-significant trend |
| ≥270,000 | 6.93 | (-0.42, 14.28) | 0.064 | Non-significant trend |
| Missing | 5.10 | (1.71, 8.48) | 0.003 | **Significant increase** |
| **Marital Status** |  |  |  |  |
| Never Married | Reference | - | - | - |
| Married | 0.26 | (-5.87, 6.39) | 0.933 | Non-significant |
| Separated/Widowed | -5.25 | (-13.07, 2.57) | 0.188 | Non-significant |
| **Smoking** |  |  |  |  |
| No | Reference | - | - | - |
| Yes | -5.30 | (-10.04, -0.56) | 0.028 | **Significant decrease** |
| Occasionally | -12.94 | (-21.66, -4.22) | 0.004 | **Significant decrease** |
| **Alcohol** |  |  |  |  |
| No | Reference | - | - | - |
| Yes | -1.80 | (-13.33, 9.73) | 0.759 | Non-significant |
| Occasionally | 10.53 | (3.26, 17.80) | 0.005 | **Significant increase** |

**Table 8.**
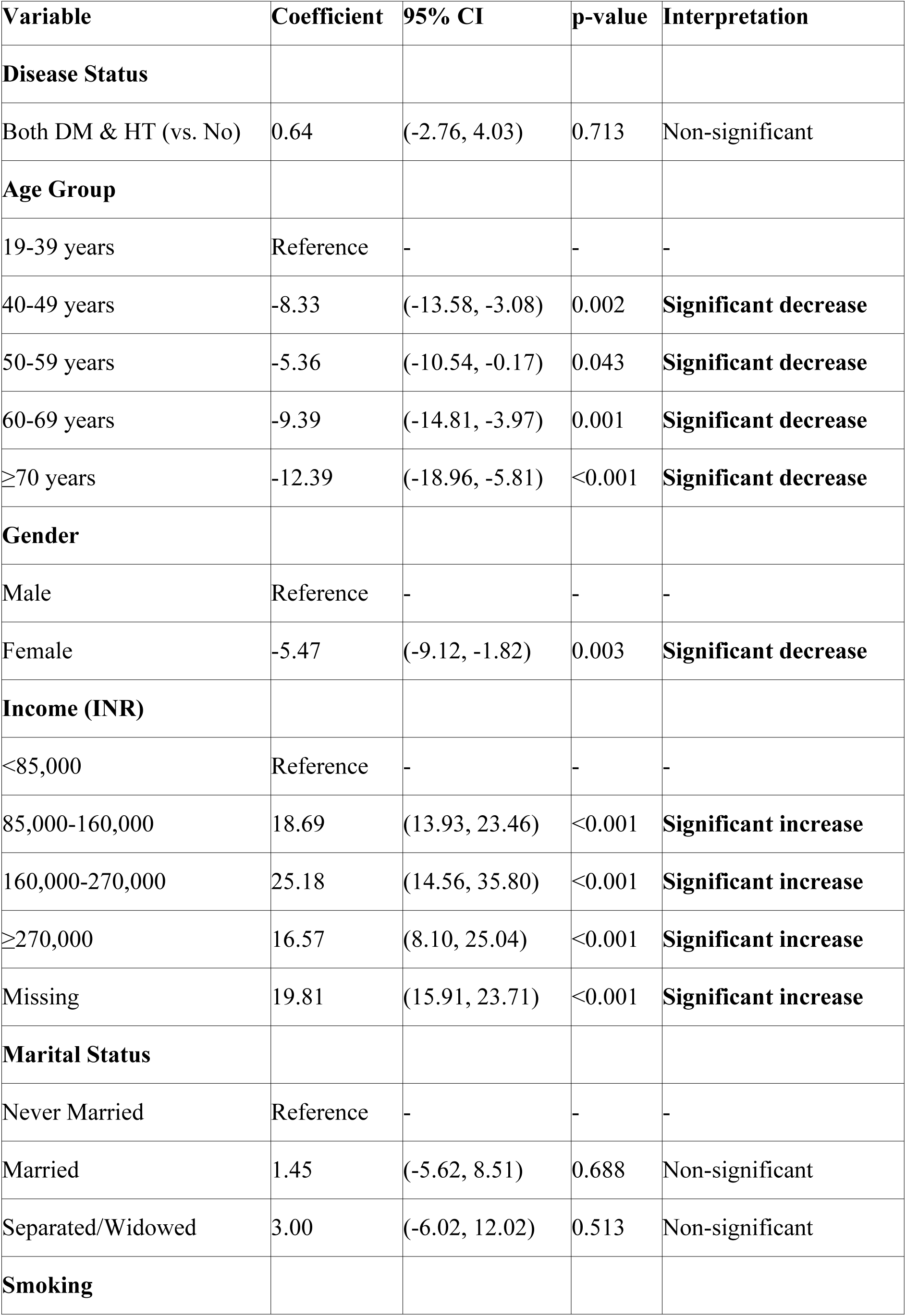
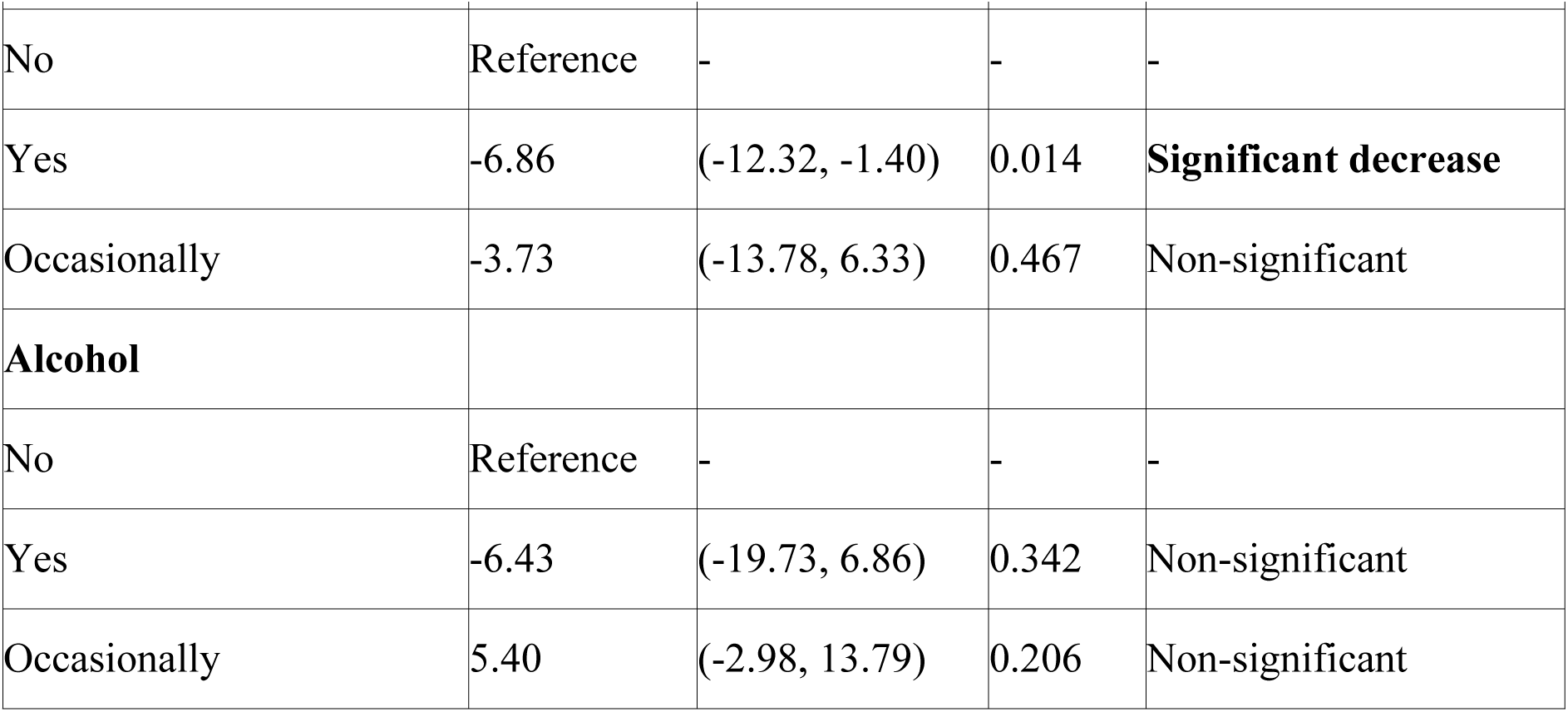
Environmental Domain Quality of Life.

**Table 9:** Summary of Significant Predictors Across Quality of Life Domains.

| Predictor | Physical Domain | Psychological Domain | Environmental Domain |
| --- | --- | --- | --- |
| **Disease Status** |  |  |  |
| Both DM & HT | ↓ (p=0.075)† | ↓ (p=0.048)* | ↔ (p=0.713) |
| **Age (vs. 19-39 years)** |  |  |  |
| 40-49 years | ↓↓ (p<0.001)* | ↓ (p=0.002)* | ↓ (p=0.002)* |
| 50-59 years | ↓↓ (p<0.001)* | ↓ (p=0.028)* | ↓ (p=0.043)* |
| 60-69 years | ↓↓↓ (p<0.001)* | ↓ (p=0.006)* | ↓ (p=0.001)* |
| ≥70 years | ↓↓↓↓ (p<0.001)* | ↓↓ (p<0.001)* | ↓↓ (p<0.001)* |
| **Gender** |  |  |  |
| Female (vs. Male) | ↓ (p<0.001)* | ↓ (p=0.001)* | ↓ (p=0.003)* |
| **Income (vs. <85,000)** |  |  |  |
| 85,000-160,000 | ↑ (p<0.001)* | ↑ (p<0.001)* | ↑↑ (p<0.001)* |
| 160,000-270,000 | ↑ (p=0.078)† | ↑ (p=0.052)† | ↑↑ (p<0.001)* |
| ≥270,000 | ↔ (p=0.132) | ↔ (p=0.064)† | ↑ (p<0.001)* |
| **Smoking** |  |  |  |
| Yes (vs. No) | ↔ (p=0.426) | ↓ (p=0.028)* | ↓ (p=0.014)* |
| Occasionally | ↔ (p=0.245) | ↓↓ (p=0.004)* | ↔ (p=0.467) |
| **Alcohol** |  |  |  |
| Occasionally (vs. No) | ↑↑ (p<0.001)* | ↑ (p=0.005)* | ↔ (p=0.206) |

\* Significant

## Discussion

This study demonstrates that QoL is significantly compromised among adults with diabetes and hypertension in urban slum primary care settings, with older adults, females, and those with uncontrolled disease and multiple medications disproportionately affected. These gradients mirror findings from other LMICs and emphasize the multidimensional nature of chronic disease burden beyond clinical indicators.

The use of the WHOQOL-BREF enabled nuanced assessment of physical, psychological, social, and environmental dimensions, revealing consistent socioeconomic disparities that highlight the need for integrated interventions addressing both medical and social determinants.

Limitations include the cross-sectional design limiting causal inference, reliance on self-reported disease control, and potential selection bias inherent in program-based recruitment.

The findings reinforce the importance of holistic primary care approaches that incorporate mental health support, patient education, and social assistance programs to enhance QoL outcomes for vulnerable urban populations. Longitudinal studies and intervention trials are needed to further elucidate drivers of QoL change and to evaluate targeted strategies.

## Conclusions

Quality of life among adults with diabetes and hypertension in urban outreach clinics is influenced by demographic, socioeconomic, and clinical factors including disease control and multimorbidity. Strengthening integrated, patient-centered chronic disease management that addresses these determinants is crucial for improving well-being and health outcomes in underserved communities.

## Data Availability

All relevant data are within the manuscript and its Supporting Information files.

## Acknowledgments

Dr Maninder Sethia, biostatistician created the analysis plan and analysed the raw data. The Health Outreach team of the Sir HN Hospital supported the research by providing the subjects from the program’s patients. Dr Charulata Pamnani and Dr Tarang Gianchandani are acknowledged for encouraging the research in the community.

The authors would like to acknowledge the Program Management team of the Health Outreach Department for supporting the research, and both Dr Charulata Pamnani and Dr Tarang Gianchandani for encouraging the authors in carrying out community research in addition to their regular work.

